# The Endothelial Dysfunction and Pyroptosis Might Drive the SARS-CoV-2 Lung Injury to the Systemic Immunothrombosis

**DOI:** 10.1101/2020.06.17.20133124

**Authors:** Seigo Nagashima, Monalisa C. Mendes, Ana Paula C. Martins, Nicolas H. Borges, Thiago M. Godoy, Anna Flavia Ribeiro dos Santos Miggiolaro, Felipe S. Dezidério, Lucia de Noronha, Cleber Machado-Souza

**Affiliations:** School of Medicine - Postgraduate Program of Health Sciences Pontifícia Universidade Católica do Paraná (PUCPR). R. Imaculada Conceição, 1155 - Prado Velho, Curitiba, PR, Brazil; Postgraduate in Biotechnology Applied in Health of Children and Adolescent. Faculdades Pequeno Príncipe (FPP). Instituto de Pesquisa Pelé Pequeno Príncipe (IPPPP). R. Silva Jardim, 1632 - Água Verde, Curitiba, PR, Brazil; Laboratory of Experimental Pathology - School of Medicine - Pontifícia Universidade Católica do Paraná (PUCPR). R. Imaculada Conceição, 1155 - Prado Velho, Curitiba, PR, Brazil; School of Medicine - Pontifícia Universidade Católica do Paraná (PUCPR). R. Imaculada Conceição, 1155 - Prado Velho, Curitiba, PR, Brazil

**Keywords:** Caspase-1, COVID-19, Endothelial dysfunction, Influenza A, Pyroptosis

## Abstract

**Objective:** Endothelial cells that are close to the alveolar-capillary exchange membranes can be activated by SARS-CoV-2 infection leading to cytokine release and macrophage activation syndrome. This could trigger endothelial dysfunction, pyroptosis, and immunothrombosis, which are the vascular changes commonly referred to as COVID-19 endotheliopathy. Thus, this study aimed to identify tissue biomarkers associated with endothelial activation/dysfunction and the pyroptosis pathway in the lung and myocardial samples of COVID-19 patients and to compare them to pandemic Influenza A virus H1N1 subtype – 2009 and Control cases.

**Approach and Results:** Post-mortem lung (COVID-19 group=6 cases; H1N1 group=10 cases, and Control group=11 cases) and myocardial samples (COVID-19=2 cases and control=1 case) were analyzed using immunohistochemistry and the following monoclonal primary antibodies: anti-CD163, anti-interleukin-6 (IL-6), anti-tumor necrosis factor-alpha (TNF-alpha), anti-intercellular adhesion molecule-1 (ICAM-1), and anti-caspase-1. From the result, IL-6, TNF-alpha, ICAM-1, and caspase-1 showed higher tissue expression in the COVID-19 group than in the H1N1 and control groups.

**Conclusion:** Our results demonstrated the presence of endotheliopathy and suggest the participation of the pyroptosis pathway in both the pulmonary and myocardial samples. These conditions might lead to systemic immunothrombotic events that could impair the efforts of clinical staff to avoid fatal outcomes. One of the goals of health professionals should be to identify the high-risk of immunothrombosis patients early to block endotheliopathy and its consequences.

## INTRODUCTION

The outbreak of the coronavirus disease 2019 (COVID-19), caused by severe acute respiratory syndrome coronavirus 2 (SARS-CoV-2), and its rapid spread have caused a global pandemic. Consequently, on January 30, 2020, the World Health Organization (WHO) declared the pandemic a Public Health Emergency of International Concern–the Organization’s highest alert level,^1,2^ and since then, the numbers of patients requiring intensive care treatment have been increasing continuously.

Following the injury in pneumocytes triggered by SARS-CoV-2 infection, an exponential release of inflammatory cytokines (cytokine storm), might be responsible for the alveolar-capillary endothelial activation. However, the endothelial cells also express ACE-2 type receptors,^3^ thus alveolar-capillary endothelial activations may be exacerbated by both the SARS-CoV-2 infection and cytokine storm, after the damage of the alveolar epithelial barrier via viral injury.^4^

The pulmonary capillary endothelium, acting as a quiescent barrier and being part of the gas exchange alveolar-capillary membrane, may switch to a responsive inflammatory phenotype after its activation, expressing cytokines and vascular adhesion molecules that could aggravate the cytokine storm,^5^ and lead to the immunothrombosis phenomena.^4,6^

Glycocalyx, a multi-component layer of proteoglycans and glycoproteins covering the luminal pole of the endothelial cells, prevents blood cell rolling and diapedesis by covering adhesion molecules (intercellular adhesion molecule-1 - ICAM-1), and participates in the regulation of the coagulation cascade. In this contest, the loss of endothelial cell glycocalyx can contribute to the breakdown of the vascular homeostasis leading to endothelial dysfunction.^7-11^

Some evidence suggests that the peculiar endothelial dysfunction induced by SARS-CoV-2 infection differs from that of the other pandemic respiratory diseases, such as pandemic Influenza A virus H1N1 subtype - 2009 (H1N1pdm09), and could link diffuse alveolar damage (DAD) with the severe systemic immunothrombosis phenomenon in patients with severe forms of COVID-19.^12^ In addition to COVID-19 endothelial activation, the significant involvement of the pyroptosis mechanism in COVID-19, but not in H1N1pdm09, may drive massive endothelial cell death thereby contributing to the thrombogenic mechanism.^13,14^

Pyroptosis is a specific type of programmed pro-inflammatory cell death^15^ that uses a cytosolic complex, called inflammasome. This structure mediates the activation of caspase-1 in response to viruses, foreign substances, and bacteria and their metabolic products.^16-18^

This article hypothesizes that, unlike the H1N1pdm09, SARS-CoV-2 endothelial activation followed by pyroptosis, may increase the pro-inflammatory stimuli, leading the COVID-19 DAD to a severe systemic immune-thrombotic disease that may impair the efforts of clinical staff to prevent fatal outcomes. Viewed from the intensive care perspective, one aim is to avoid COVID-19 systemic vascular injury as a way to prevent the almost always lethal immune-thrombotic phenomenon.

This study aimed to analyze endothelial activation and pyroptosis pathway tissue biomarkers in patients who died from COVID-19 compared with cases of H1N1pdm09 deaths and controls.

## METHODS

### Study population

The COVID-19 group comprised lung samples from post-mortem biopsies of patients whose cause of death was SARS-CoV-2-induced diffuse alveolar damage during the COVID- 2019 pandemic. In patients 4 and 6, in addition to the lung, there were myocardial samples. The clinical data of the six cases were obtained from their medical records during hospitalization in the Intensive Care Unit (ICU) at Hospital Marcelino Champagnat in Curitiba, Brazil. The COVID-19 group (n=6) was approved by the National Research Ethics Committee (REC: 3.944.734/2020).

The H1N1 group (n=10) comprised lung samples from post-mortem biopsies of patients whose cause of death was H1N1pdm09 severe acute respiratory infections during the 2009 pandemic. The clinical data of all the cases were obtained from their medical records during hospitalization in the ICU at Hospital de Clínicas in Curitiba, Brazil (REC: 2.550.445/2018).

The control group (n=12) comprised of lung (n=11) and myocardial (n=1) samples from necropsies of patients who died from other causes, not involving lung and myocardial lesions. The clinical data of all the cases were obtained from their medical records during hospitalization in the ICU at Hospital de Clínicas in Curitiba, Brazil (REC: 2.550.445/2018).

Testing for H1N1pdm09 and SARS-CoV-2 was performed on nasopharyngeal swabs taken during ICU hospitalization, and real-time reverse transcriptase-polymerase chain reaction (rRT-PCR) was positive in all cases. The families of the patients consented to the post-mortem biopsy for the cases of COVID-19, H1N1 and Control groups.

### Immunohistochemical assays

Minimally invasive lung/myocardial biopsies were performed through a left anterior mini-thoracotomy with upper left lobe lingular segment and left ventricular wall fragment resection. The resected pieces were 3×3 cm.

The lung and heart samples provided by post-mortem biopsies/necropsies of all three groups were formalin-fixed paraffin-embedded (FFPE) and stained with hematoxylin and eosin (H&E) to find the appropriate areas for the immunohistochemical techniques. The FFPE lung/heart samples were also performed using immunohistochemical reactions and the primary monoclonal antibodies, anti-CD163 (rabbit polyclonal, clone 14215, Thermo Fisher Scientific, 1:1000), IL-6 (mouse monoclonal, clone ab9324, Abcam, 1:400), TNF-alpha (rabbit polyclonal, clone ab6671, Abcam, 1:100), ICAM-1 (mouse monoclonal, clone 23G12, Novocastra, 1:100), and caspase-1 (rabbit polyclonal, ab189796, Abcam, 1:200).^19^

The macrophages score was performed by scoring CD163+ cells in 30 high-power fields (HPFs). The HPFs were chosen randomly from the septum and lumen alveolar.

The lung tissue immune slides of IL-6, TNF-alpha, and ICAM-1 were scanned on the Axio Scan Scanner Z1 (Carl Zeiss, Germany). Afterward, 10 HPFs per case were selected from the area of interest (vessels). The measurement was made only in the vascular endothelium, and the positive areas appeared through a semi-automatic segmentation method of quantification, using the Image-Pro Plus software version 4.5 (Media Cybernetics, USA). Subsequently, these areas were converted into percentages to enable statistical analysis.

A semiquantitative analysis was performed for caspase-1 using the Allred score in the endothelial cells of the lung samples. It was obtained by summing two scores (proportion and intensity of positivity) ranging from 0–8. The proportion score was subdivided according to the percentage of stained cells: score 0 (0% stained cells), score 1 (<1%), score 2 (1–10%), score 3 (11–33%), score 4 (34–66%), and score 5 (> 66%). The intensity of positivity was evaluated as follows: negative - score 0, weak - score 1, moderate - score 2, and strong - score 3.^20^ The scoring was performed taking into account only the vascular endothelium.

The three myocardium immune slides (cases 4 and 6 of the COVID-19 group and one control patient) were also analyzed using the Allred score for CD163 (macrophages score), IL-6, TNF-alpha, ICAM-1, and caspase-1.

### Statistical analysis

Descriptive analysis was performed using the absolute number and relative frequencies of the qualitative variables. The quantitative variables were described by means and standard deviation, and by medians with minimum and maximum values. The normality condition of the variables in each group was evaluated using the Shapiro-Wilks test. The Kruskal-Wallis test was used for comparison of the quantitative variables. Data were analyzed using the computer program R Project for Statistical Computing. Data were analyzed using the IBM® SPSS Statistics v.20.0 software. Values of *p* < 0.05 were considered statistically significant.

## RESULTS

The baseline characteristics of COVID-19 patients are presented in Table 1. The lung sample immunohistochemical results in all groups are presented in Table 1 and Figure 1.

**Table 1.** Tabular comparison between COVID-19, H1N1, and Control groups for immunohistochemical expression of endothelial dysfunction biomarkers in the lung and myocardial tissues.

| Samples | Variables | COVID-19 (n=6) | H1N1 (n=10) | p-value* | Control (n=11) | p-value† |
| --- | --- | --- | --- | --- | --- | --- |
| Lung | CD163<sup>1,2,3</sup> | 52.9 (14.4-87.8)<sup>1</sup> | 71.4 (37.1-71.4) | 0.1931 | 31.5 (19.5-47.6) | 0.3149 |
|  |  | 49.0±28.7<sup>2</sup> | 69.2±20.3 |  | 31.7±8.2 |  |
|  | IL-6<sup>1,2,4</sup> | 3.9 (0.3-7.6) | 1.3 (0.7-3.1) | <u>0.0652</u> | 0.0 (0.0-5.6) | <u>0.0049</u> |
|  |  | 4.0±2.5 | 1.6±0.9 |  | 0.6±1.6 |  |
|  | TNF-alpha<sup>1,2,4</sup> | 5.8 (3.2-35.3) | 2.1 (0.1-4.9) | <u>0.0092</u> | 2.2 (0.2-4.1) | <u>0.0048</u> |
|  |  | 12.3±12.9 | 2.5±1.5 |  | 1.9±1.5 |  |
|  | ICAM-1<sup>1,2,4</sup> | 4.3 (1.4-20.6) | 0.9 (0.0-3.3) | <u>0.0034</u> | 0.4 (0.1-2.6) | <u>0.0034</u> |
|  |  | 7.4±7.3 | 1.0±1.0 |  | 0.8±0.8 |  |
|  | Caspase-1<sup>1,2,5</sup> | 8.0 (7.2-8.0) | 4.5 (3.0-7.0) | <u>0.0009</u> | 2.0 (2.0-4.0) | <u>0.0009</u> |
|  |  | 7.8±0.3 | 4.8±1.2 |  | 2.4±0.7 |  |
| Myocardial | CD163<sup>6</sup> | 7.0<sup>‡</sup> | -- | -- | 5.0<sup>§</sup> | -- |
|  | IL-6<sup>6</sup> | 8.0<sup>‡</sup> | -- | -- | 5.0<sup>§</sup> | -- |
|  | TNF-alpha<sup>6</sup> | 7.0<sup>‡</sup> | -- | -- | 6.0<sup>§</sup> | -- |
|  | ICAM-1<sup>6</sup> | 8.0<sup>‡</sup> | -- | -- | 3.0<sup>§</sup> | -- |
|  | Caspase-1<sup>6</sup> | 8.0<sup>‡</sup> | -- | -- | 3.0<sup>§</sup> | -- |

Legends: <sup>1</sup>Median (Min-Max). <sup>2</sup>mean±standard deviation. <sup>3</sup>mean number of CD163+ macrophages in 30 high power fields (HPF). <sup>4</sup>IL-6/TNF-alpha/ICAM-1 tissue expression in percentage per HPF. <sup>5</sup>Allred score for caspase-1 in lung samples. \*p-values obtained were compared between COVID-19 versus H1N1. †p-values obtained were compared between the COVID-19 and Control group. <sup>6</sup>Allred score in myocardial samples. ‡Myocardial samples of the COVID-19 patients 4 and 6. §Control patient myocardial sample: female, 80 years old; hypertension and pulmonary thromboembolism after left knee arthroplasty. p-values were determined using the non-parametric Kruskal-Wallis test.

**Figure 1.**
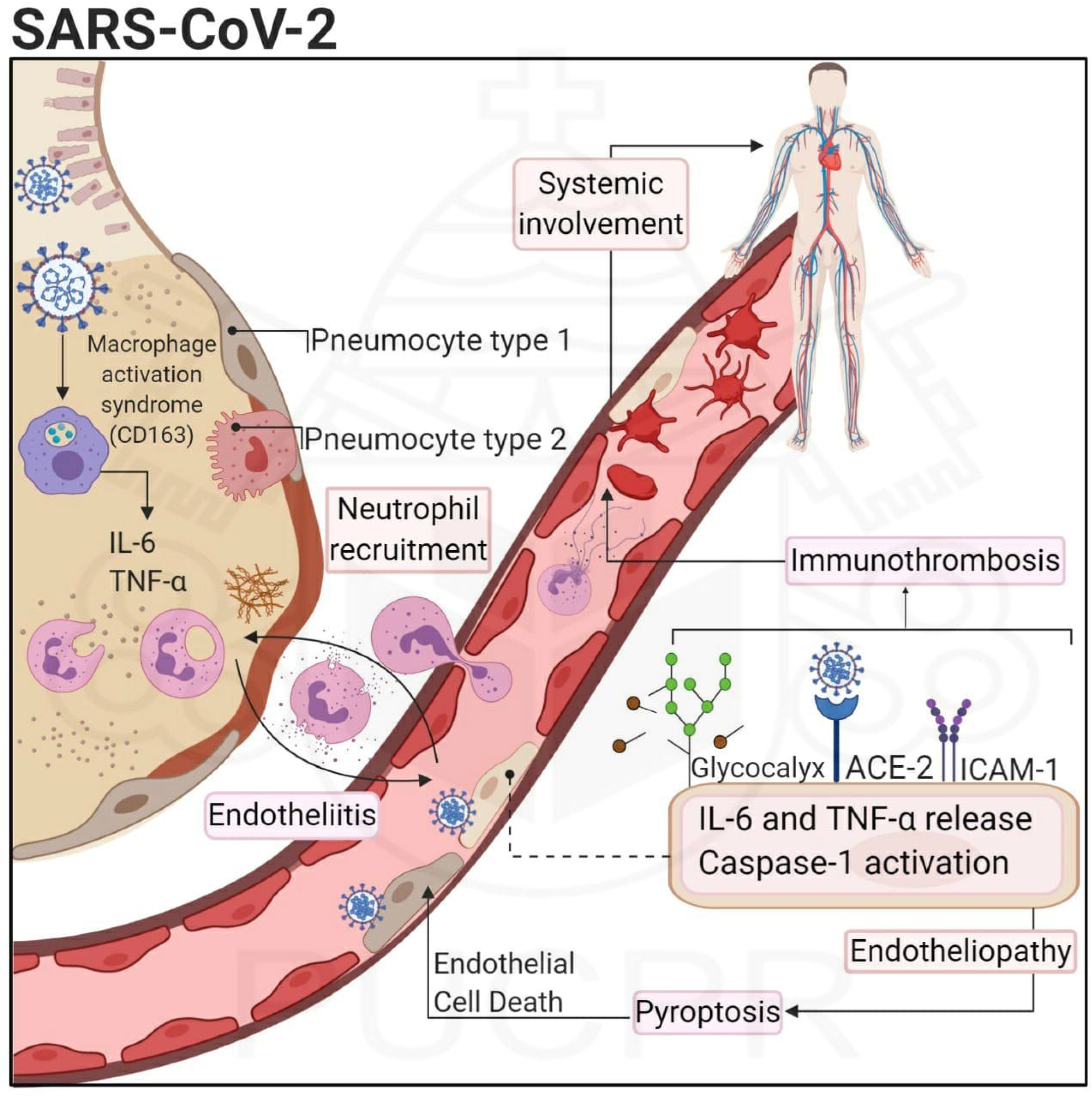
Graphs showing the comparison between the COVID-19 and H1N1 groups regarding TNF-alpha, IL-6, ICAM-1 (in percentage per HPF), and caspase-1 tissue expression (in Allred score). TNF-alpha is remarkably high in the alveolar septal cells (black arrowheads) and the alveolar-capillary cells (black arrows) of the COVID-19 case than in the H1N1 case. Alveolar lumens are identified with asterisks. IL-6 tissue expression is remarkably high in the endothelial cells (black arrows) of the COVID-19 case than in the H1N1 case. ICAM-1 tissue expression is remarkably higher in the endothelial cells (black arrows) of the COVID-19 case than in the H1N1 case. Caspase-1 is higher in the endothelial cells of the COVID-19 case than in the H1N1 case. Kruskal-Wallis test was used for *p-* values. Scale bars = 50 micrometers.

The demographic data were analyzed and presented using the mean and standard deviation. Comparing the biological sex between the COVID-19 group (male n=4, 66.6%, female n=2, 33.4%) versus the H1N1 group (male n=8, 80.0%; female n=2, 20.0%), and found no significant difference (*p*=0.5510). Similarly, there was no significant difference (*p*=0.7933) between the COVID-19 and Control group (male n=8, 72.7%; female n=3, 27.3%). The mean age was significantly (*p*=0.0040) higher in the COVID-19 group (76.5±12.5) than in the H1N1 group (43.5±13.9). Similarly, the mean age was significantly (*p*=0.0020) higher in the COVID-19 group than in the Control group (42.3±14.3). Survival days in COVID-19 group (18.8±11.6) were significantly (*p*=0.0058) higher than in the H1N1 group (4.7±6.1). Comparably, significant differences (*p*=0.0109) were observed between the COVID-19 and Control groups (7.6±13.1). Mechanical ventilation was not significantly (*p*=0.1851) different between the COVID-19 (9.7±6.8) and the H1N1 group (4.7±6.1).

The histopathological analysis with H&E staining is presented in Supplementary Figure I. The COVID-19 and H1N1 lung samples presented DAD. There were slight number of septal and intra-alveolar neutrophils, and numerous small fibrinous thrombi were observed in the small and medium pulmonary arteries followed by neutrophilic endotheliitis and endothelial cell tumefaction in the COVID-19 group. However, there was no significant endothelial activation, fibrinous thrombi, and neutrophilic endotheliitis in the H1N1 cases.

Comparatively, IL-6 (*p*=0.0652), TNF-alpha (*p*=0.0092), ICAM-1, and caspase-1 (*p*=0.0009) were highly expressed in the lung tissues in the COVID-19 group than in the tissues in the H1N1 group (Figure 1). The same pattern was observed in the COVID-19 compared with the Control group (Table 1).

The immunohistochemical results of myocardial post-mortem biopsies/necropsies are summarized in Table 1.

Supplementary Figure I shows the immunohistochemical results of the COVID-19 myocardial samples (n=2).

## DISCUSSION

This study aimed to understand the context of the endothelial activation and pyroptosis in patients that died of COVID-19 after hospitalization, as these events may have a possible influence on subsequent thrombogenic events.

Endothelial activation refers to the ability of the endothelium to regulate vascular homeostasis and mainly describes its imbalance in favor of the pro-inflammatory and prothrombotic effects.^21^ Pyroptosis increases the release of pro-inflammatory cytokines such as IL-1 beta and IL-18.^22^ Infection by SARS-CoV-2 would possibly contribute to endothelial dysfunction and to the peculiar endothelial cell death called pyroptosis.^23^ Thus, these two events could also trigger the subsequent immunothrombotic process that could have a significant impact on the thrombogenic mortality observed in COVID-19 patients.

The endothelium, without this activation process, provides a suitable luminal surface that does not promote the activation of the intrinsic coagulation cascade or platelet adhesion, but anticoagulant and fibrinolytic mechanisms.^24^ The capillary-alveolar endothelial cells receive a “shower of cytokines” (IL-1, IL-6, and TNF-alpha) from the pneumocytes infected by SARS-CoV-2. After that, the endothelial cells are activated and they produce NF-κB and adhesion molecules such as ICAM-1.^25-27^ Thus, under these inflammatory conditions, a subset of leukocytes (polymorphonuclear leukocytes, macrophages, and mast cells) degranulate enzymes that can contribute to the breakdown of the glycocalyx of the endothelial cells.^28,29^ The intact glycocalyx serves as a barrier against the platelets and leukocytes, and the impairment of its function would help lead to endotheliitis and the thrombotic events.^30-34^

The macrophages could amplify the local pro-inflammatory response by an early recruitment of the complement system.^35^ Recent reports suggest that the cytokine storm caused by SARS-CoV-2 infection has significant similarities to the findings observed in macrophage activation syndrome.^36^ This syndrome is associated with high secretion of several cytokines, including IFN-gamma, IL-1, IL-6, IL-18, and TNF-alpha.^37,38^ The observation of the high number of macrophages (CD163 score) in the COVID-19 group might suggest the involvement of these cells in the mechanism of COVID-19.

Following this pro-inflammatory pattern, the higher and continuous tissue expression of IL-6 and TNF-alpha in the COVID-19 group than in the H1N1 and Control groups (*p*=0.0652-borderline and *p*=0.0049; *p*=0.0092 and *p*=0.0048, respectively) demonstrate that inflammatory response may be kept active throughout the evolvement of COVID-19. It is known that the cytokine profile of patients with COVID-19 showed a marked increase in interferon-gamma, IL-1, IL-6, and IL-12, at least two weeks after the onset of the disease.^39-41^

Comparatively, the expression of ICAM-1 in the tissues was significantly higher in the COVID-19 group (*p*=0.0034) than in the H1N1 or Control groups. An essential consequence of endothelial activation is the expression of adhesion molecules such as the ICAM family.

These molecules have the classic function of attracting leukocytes to the infected region and have the ability to transmit intracellular signals leading and keeping the pro-inflammatory status, thus perpetuating the innate immune response and subsequently promoting the systemic activation of the endothelial cells.^42,43^ The continuous pro-inflammatory condition would result not only in a dysfunctional endothelium but also lead to the loss of its integrity via endothelial cell death.^44^ The persistent inflammatory signaling of these adhesion molecules would also contribute to later thrombogenic effects. Patients 4 and 5 (respectively 38 and 23 days of survival) showed the highest vascular tissue expression of ICAM-1, and this observation could be associated with severe thrombotic events that were observed in these cases.

The relationship between endothelial activation/dysfunction and subsequent thrombotic events is already well-known in cardiovascular diseases^45^ and diabetes.^46,47^ Five of our cases had comorbidities (data not shown), such as systemic arterial hypertension, dyslipidemia, diabetes, chronic kidney disease, and signs of chronic arterial disease. Thus, the permanent status of endothelial activation promoted by these comorbidities would aggravate endothelial dysfunction caused by a viral inflammatory response and could be responsible for the most prevalent fatal outcome described in these patients.^48^ Suo and colleagues^49^ showed that H1N1pdm09 is responsible for potentiating endothelial cell apoptosis in patients who already have atherosclerosis. In this context, SARS-CoV-2 infection of the pneumocytes, besides having an essential role in the inflammatory activation/dysfunction of endothelial cells, would also lead to endothelial cell death, causing the discontinuity of the alveolar-capillary barrier and facilitating the movement of the virus from the alveolar septum through the alveolar-capillary lumen. Thus, endothelial cells would have their glycocalyx layer damaged by leukocytes and would become a further target for SARS-CoV-2 by binding the ACE-2 receptor, exposed in the endothelial cell membrane.

Pyroptosis is an inflammatory form of programmed cell death,^50^ and occurs by activating a protein complex known as inflammasome that is formed by the union of several molecules (NLRP3, ASC, and procaspase-1).^22,23,51,52^ Caspase-1 produces morphological changes through the creation of membrane pores,^53^ cell edema, the release of pro-inflammatory cytokines (IL-1 beta and IL-18), and cell fragmentation.^54^ This final event resulting from pyroptosis would prevent the replication of pathogens,^55-57^ and the interleukin release would alert the immune system. It has been proposed that pyroptosis could contribute to the death of endothelial cells after SARS-CoV-2 infection.^23^ A group of researchers found that SARS-CoV-2 proteins can trigger the mechanism of cellular pyroptosis.^22^ In another study, Middle East respiratory syndrome coronavirus (MERS-CoV)-infected macrophages promote high pyroptotic biomarker expression.^58^ Our results showed higher endothelial tissue expression of caspase-1 in the COVID-19 group than in the H1N1 and Control groups (*p*=0.0009). Only the presence of high levels of caspase-1 does not confirm the occurrence of the pyroptosis process. However, the remarkable presence of one marker of pyroptosis (caspase-1) is indicative of a probable association of SARS-CoV-2 endothelial infection with this type of inflammatory cell death in endothelial cells.

The presence of a similar pattern of tissue expression in the myocardial samples (higher tissue expression of CD163 score, IL-6, ICAM-1, TNF-alpha, and Caspase-1 in COVID-19 patients than in the control patients) might suggest a systemic involvement. However, the authors understand that the small number of myocardial samples (n=2) only allows us to conjecture this hypothesis, and further studies would be necessary to prove it.

The main limitations of this study are the small number of cases (n=6)^4^ and data based on FFPE post-mortem samples that can only provide a piece of static information at the time of the death and cannot reconstruct the evolving disease process. A limitation that might be influencing the expression of biomarkers would be the non-pairing baseline characteristics in the three groups (age and survival times). However, the COVID-19 and H1N1 groups are very different demographically from each other because these two pandemic diseases have marked differences in their risk groups and pathophysiology. In addition, the survival time is quite different between these two groups, since patients in the H1N1 group died earlier than has been observed in patients with COVID-19. We paired these two groups by mechanical ventilation time to minimize the differences in inflammatory response caused by this type of intervention.

Our results suggest that endothelial activation/dysfunction and pyroptosis, also called endotheliopathy,^59^ could lead to a systemic immunothrombotic disease. It is known that thrombogenic conditions are quite characteristic in COVID-19^3^ patients. Understanding the complex events that can be part of an immunothrombosis mechanism might help the health professional identify early patients with the potential of endotheliopathy based on predisposing comorbidities (chronic renal disease, diabetes, and hypertension) and systemic biomarkers, such as D-dimer, angiopoitin-2, and thrombomodulin.^59,60^ The early diagnosis of endotheliopathy could guide health professionals in implementing therapeutic anticoagulation and endothelial stabilization in high-risk patients, mitigating thrombogenic events, systemic inflammatory response syndrome (SIRS), and multiple organ failure.

In conclusion, our findings lead us to suggest the involvement of endotheliopathy, pyroptosis, and immunothrombosis in COVID-19. This immunothrombosis event may be responsible for the devastating clinical consequences with fatal outcomes in the severe form of this pandemic disease COVID-19.

## Data Availability

The authors confirm that all data referred in the manuscript are available.

## Acknowledgments

We are grateful to the Universidade Federal do Paraná, and Pontificia Universidade Católica do Paraná for their support.

## Nonstandard abbreviations and acronyms

TNF-alpha: tumor necrosis factor-alpha
IL: interleukin
H1N1pdm09: pandemic Influenza A virus H1N1 subtype disease 2009
ICAM-1: intercellular adhesion molecule 1
DAD: diffuse alveolar damage
HPF: high-power field
MAS: macrophage activation syndrome
SARS-CoV-2: severe acute respiratory syndrome coronavirus 2
COVID-19: pandemic coronavirus disease 2019
NLRP3: NOD-like receptors protein 3
NF-kB: nuclear factor-kappa-B
SIRS: systemic inflammatory response syndrome

## Highlights

- COVID-19 post-mortem lung and myocardial samples showed higher endothelial expression of IL-6, TNF-alpha, ICAM-1, and caspase-1 than the H1N1 and Control samples.
- There is some evidence of pyroptosis and endotheliopathy in the COVID-19 lung and myocardial samples compared to the H1N1 and Control samples.
- Endotheliopathy and pyroptosis can lead to systemic immunothrombosis that could impair the efforts of clinical staff to prevent fatal outcomes.
- COVID-19 patients could benefit if the high risk of immunothrombosis is identified early to block the endotheliopathy and its consequences.

## Legends

**Graphical Abstract:** SARS-CoV-2 infection followed by macrophage activation (MAS), the release of cytokines, neutrophil recruitment, endotheliopathy, and pyroptosis, promoting immunothrombosis that leads to systemic disease.

---

**Supplementary Figure I.**
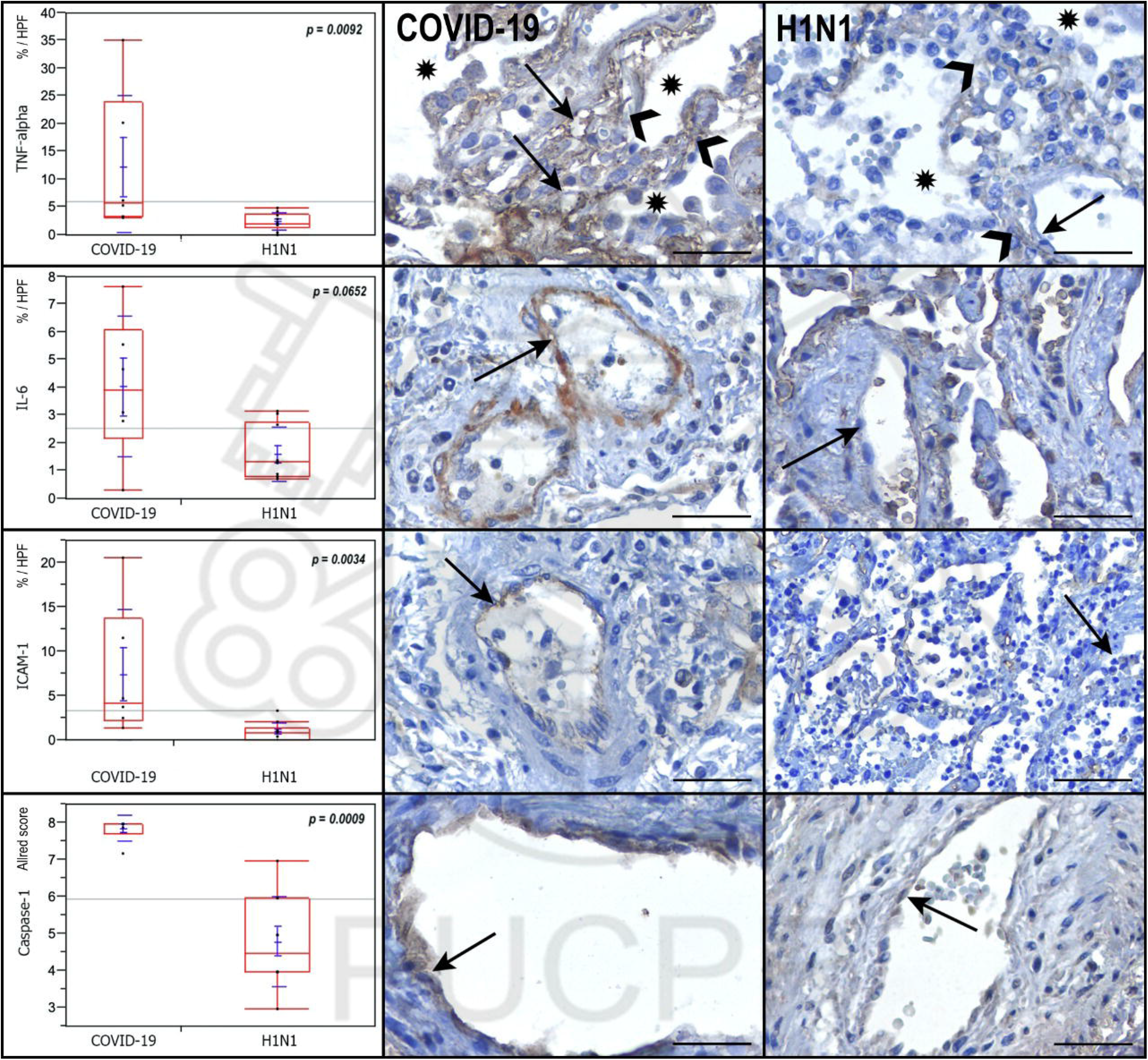
COVID-19 myocardial sample (case 6) expressing caspase-1 in the endothelium (black arrows). However, the control case did not express caspase-1 in the endothelial cells (black arrow). ICAM-1 expression in the endothelial cells (black arrows) in the COVID-19 myocardial sample (case 6) is more pronounced than that in the control case (black arrow). Endothelial cells and macrophages strongly expressed TNF-alpha (black arrows) in the COVID-19 myocardial sample (case 4) than in the control case, where TNF-alpha is expressed only in the macrophages (black arrow). In the COVID-19 case 3, H&E conventional staining technique shows vessel (asterisk) with endothelial cell tumefaction and endotheliitis (left) and diffuse alveolar damage (DAD) characterized by thickened septa and hyaline membranes (arrowheads).

